# Pathway-Specific Polygenic Scores Improve Cross-Ancestry Prediction of Psychosis and Clinical Outcomes

**DOI:** 10.1101/2023.09.01.23294957

**Authors:** Justin D. Tubbs, Perry B.M. Leung, Yuanxin Zhong, Na Zhan, Tomy C.K. Hui, Karen K.Y. Ho, Karen S.Y. Hung, Eric F.C. Cheung, Hon-Cheong So, Simon S.Y. Lui, Pak C. Sham

**Affiliations:** Psychiatric and Neurodevelopmental Genetics Unit, Center for Genomic Medicine, Massachusetts General Hospital, Boston, MA, USA; Department of Psychiatry, Harvard Medical School, Boston, MA, USA; Stanley Center for Psychiatric Research, Broad Institute of MIT and Harvard, Cambridge, MA, USA; Department of Psychiatry, Li Ka Shing Faculty of Medicine, The University of Hong Kong, Pok Fu Lam, Hong Kong SAR; Department of Psychosis Studies, Institute of Psychiatry, Psychology and Neuroscience, King’s College London, London, UK; Department of General Adult Psychiatry, Castle Peak Hospital, Hong Kong SAR; School of Biomedical Sciences, The Chinese University of Hong Kong, Shatin, Hong Kong SAR; Department of Psychiatry, The Chinese University of Hong Kong, Shatin, Hong Kong SAR; Centre for PanorOmic Sciences, Li Ka Shing Faculty of Medicine, The University of Hong Kong, Hong Kong SAR; State Key Laboratory of Brain and Cognitive Sciences, The University of Hong Kong, Hong Kong SAR

## Abstract

Psychotic disorders are debilitating conditions with disproportionately high public health burden. Genetic studies indicate high heritability, but current polygenic scores (PGS) account for only a fraction of variance in psychosis risk. PGS often show poor portability across ancestries, performing significantly worse in non-European populations. Pathway-specific PGS (pPGS), which restrict PGS to genomic locations within distinct biological units, could lead to increased mechanistic understanding of pathways that lead to risk and improve cross-ancestry prediction by reducing noise in genetic predictors. This study examined the predictive power of genome-wide PGS and nine pathway-specific pPGS in a unique Chinese-ancestry sample of deeply-phenotyped psychosis patients and non-psychiatric controls. We found strong evidence for the involvement of schizophrenia-associated risk variants within “nervous system development” (p=2.5e-4) and “regulation of neuron differentiation” pathways (p=3.0e-4) in predicting risk for psychosis. We also found the “ion channel complex” pPGS, with weights derived from GWAS of bipolar disorder, to be strongly associated with the number of inpatient psychiatry admissions a patient experiences (p=1.5e-3) and account for a majority of the signal in the overall bipolar PGS. Importantly, although the schizophrenia genome-wide PGS alone explained only 3.7% of the variance in liability to psychosis in this Chinese ancestry sample, the addition of the schizophrenia-weighted pPGS for “nervous system development” and “regulation of neuron differentiation” increased the variance explained to 6.9%, which is on-par with the predictive power of PGS in European ancestry samples. Thus, not only can pPGS provide greater insight into mechanisms underlying genetic risk for disease and clinical outcomes, but may also improve cross-ancestry risk prediction accuracy.

## Introduction

Psychosis-related disorders including schizophrenia and bipolar disorder are chronic and recurrent conditions with a lifetime prevalence of 1-3% (Chang et al., 2017; Global Burden of Disease Collaborative Network, 2020; Huang et al., 2019; Perälä et al., 2007; Subramaniam et al., 2021), causing severe impairment in daily functioning, increased mortality (Walker et al., 2015), significant burden on caregivers (Flyckt et al., 2015; Rössler et al., 2005), and disproportionately high economic costs (Fineberg et al., 2013). Psychotic disorders are complex phenotypes with high heritabilities (Cardno et al., 2002, 1999; Gordovez and McMahon, 2020; Hilker et al., 2018; Rijsdijk et al., 2011, 2011; Sullivan et al., 2003), including significant contributions from common single nucleotide polymorphisms (SNPs) captured by genotyping arrays (Mullins et al., 2021; Trubetskoy et al., 2022).

Genome-wide association studies (GWASs) conducted by international consortia have been successful in identifying risk variants, genes, biological pathways, and brain regions associated with risk for psychotic disorders (Mullins et al., 2021; Trubetskoy et al., 2022). While these risk variants separately have a small effect size, they explain notable proportions of variance in liability to psychotic disorders when aggregated into genome-wide polygenic scores (PGS) (Mullins et al., 2021; Trubetskoy et al., 2022). However, the vast majority of GWAS participants are of western European ancestry, with current PGS methods showing relatively poor portability across ancestries for most disorders, including schizophrenia (Ju et al., 2022; Kim et al., 2018; Lam et al., 2019; Martin et al., 2019). This is despite the fact that comparative analyses of European and East Asian GWAS results indicate significant cross-ancestry genetic correlations and a high degree of overlap in enriched pathways (Lam et al., 2019).

PGS are traditionally designed to summarize individual-level disease risk using variants across the entire genome into a single number, and several previous studies have shown significant association between PGS and risk for psychotic disorders, mostly in samples of European ancestry (Mullins et al., 2021; Trubetskoy et al., 2022). However, while utilizing information from all genomic variants may optimize predictive power, it obscures individual differences in potential biological mechanisms. Pathway-specific PGS (pPGS) is a recently introduced method which attempts to re-incorporate biological information in a PGS framework by restricting variants contributing to a given pPGS to those within genes belonging to a single pathway (Choi et al., 2023). This allows explicit testing of the association between genetic variants in a specific biological pathway and risk for disease or other phenotypes, while accounting for residual individual variation in genetic risk. The possibility of constructing highly predictive pPGS has been greatly boosted by enrichment analyses of large-scale GWAS summary statistics for schizophrenia demonstrating the involvement of specific biological processes (Trubetskoy et al., 2022).

A few studies have used pPGS to predict brain endophenotypes or cognitive deficits in schizophrenia patients of European ancestry (Cosgrove et al., 2018, 2017). One study used a pPGS to predict schizophrenia case-control status in a Chinese ancestry sample (Yao et al., 2021), while one recent study has examined pPGS prediction of secondary clinical outcomes in an ancestrally-diverse sample of psychosis patients (Warren et al., 2023). We are unaware of any studies testing for association between multiple pPGS and secondary outcomes in psychosis patients in the Chinese population, which is important for understanding whether pathways conferring genetic risk for psychosis replicate in diverse global populations and how these pathways influence meaningful clinical outcomes.

Thus, the current study examines the predictive power of genome-wide PGS and nine pathway-specific pPGS in a unique Chinese-ancestry sample of deeply-phenotyped psychosis patients and non-psychiatric controls. Our main analyses aim to test whether pPGS are useful for predicting psychosis in a Chinese ancestry sample, determine whether pPGS contribute additional predictive power over genome-wide PGS, and identify the relative importance of each pathway in conferring risk for psychosis. Secondary analyses seek to understand the role of pPGS and genome-wide PGS in predicting clinical outcomes among patients.

## Methods

### Participants

Clinical participants were recruited through a research-orientated clinical programme at Castle Peak Hospital in Hong Kong (Lui et al., 2011). Patients attending the early psychosis intervention clinic between 2011 and 2015 who were diagnosed with schizophrenia or psychosis-related disorders using criteria from the DSM-IV (American Psychiatric Association, 1994) were invited to participate in the research study. Healthy controls were recruited from neighbouring community, and received structured interview with qualified psychiatrists to ensure that they did not have personal and family history of psychotic disorders. All participants provided signed written informed consent. This study has been approved by the New Territories West Cluster Research Ethics Committee (Protocol number: NTWC/CREC/823/10; NTWC/CREC/1293/14; UW14-325). The participants of this study have completed additional assessments (including endophenotyping using behavioural and neuroimaging measures), and the findings have been reported elsewhere (Cheung et al., 2015; Chiu et al., 2018; Deng et al., 2019; Lui et al., 2022, 2021, 2016, 2015).

Clinical diagnoses were ascertained based on the “best-estimate” approach, using structured interview by qualified psychiatrists, supplemented with medical records. The diagnostic composition of our sample is shown in Supplementary Table 1. From 2020-2021, we conducted a comprehensive review of patients’ computerized medical records using a proforma to retrieve the following variables: onset age, history of self-harm/suicide behaviors, history of aggressive acts, and dates of inpatient psychiatric admission and discharge. From the inpatient data, we subsequently calculated the number of distinct admission events, as well as the total length (days) of all psychiatric inpatient stays for each patient. The operational definitions of these variables are shown in Supplementary Table 2. The variables were compiled mainly from the information recorded in the computerized electronic medical record system of the Hospital Authority of Hong Kong by trained research assistants, under the supervision of qualified psychiatrists. The staff responsible for collecting the clinical phenotype variables were not involved in the data analysis in this study.

### Genetic Data Quality Control

We collected DNA samples from whole blood and performed array genotyping using the Illumina Asian Screening Array. Standard quality control was performed on the genotype data including removing samples with heterozygosity rate outside 3 standard deviations from the mean, genotype missingness rate greater than 5%, or having a mismatch between self-reported and genotype-inferred sex. Variants were filtered out if their missingness rate was greater than 5%, if missingness differed significantly between cases and controls, or if genotype frequencies differed significantly from Hardy-Weinberg equilibrium. Samples were then submitted to the Michigan Imputation Server using the 1000 genomes phase-3-v5 release as the reference panel. Post-imputation, variants were further filtered to have an imputation quality score greater than 0.8. We also filtered out one individual from any pair related closer than the 3^rd^ degree, with preference for retaining psychosis cases over controls. Genetic principal components (PCs) were fitted in the 1000 genomes East Asian subset and projected onto the current sample to adjust for population stratification in subsequent models.

### Polygenic Score Calculation

A total of 30 polygenic scores were calculated for each individual in the target sample. One overall genome-wide PGS and nine pathway-specific pPGS were calculated using the latest GWAS for schizophrenia (SCZ; Trubetskoy et al., 2022), bipolar disorder (BIP; Mullins et al., 2021), and depression (DEP; Howard et al., 2019) as training datasets. There was no overlap between participants in any of the training GWAS and the target sample.

Overall genome-wide PGS for SCZ, BIP, and DEP were calculated using PLINK v1.90b5.3 (Chang et al., 2015) with weights generated by applying PRS-cs (Ge et al., 2019) to their respective summary statistics with the phi shrinkage parameter set to 0.01 and the 1000 genomes European subset used as the LD reference panel. In addition to the overall PGS, we calculated pPGS for the nine gene ontology pathways which represented independent enrichment signals in the most recent GWAS of SCZ (Trubetskoy et al., 2022): axon, ion channel complex, nervous system development, neuronal cell body, somatodendritic compartment, synapse, regulation of cation channel activity, regulation of neuron differentiation, voltage-gated calcium channel activity. In order to ensure complete coverage of pathway genes, a clumping and thresholding approach implemented in the PRSet software package (Choi et al., 2023) was used to calculate pPGS for the nine biological pathways of interest. For pPGS calculation, the 1000 genomes European reference sample was used for LD calculation and the default PRSet parameters were used, while setting the proxy argument to 0.8, which allows strongly linked SNPs to represent genes when constructing pPGS. All PGS were mean-centered and standardized to unit variance prior to modeling.

### Modeling Approach

Our primary analysis tested whether psychosis case-control status was associated with genome-wide PGS and nine pathway-specific PGS, each calculated using weights based on SCZ, BIP, and DEP summary statistics. For each of the 30 PGS, we fit logistic regression models predicting psychosis case-control status independently for each of the 3 genome-wide PGS (3 sets of weights) and 27 pPGS (9 pathways x 3 sets of weights). For all models, we included age, sex, and the first ten genetic principal components (PCs) as covariates.

As a secondary analysis, we tested whether these genome-wide or pathway PGS were associated with each of five additional clinical outcomes of interest: onset age, self-harm/suicide, aggressive acts, total admission length, and total inpatient admissions. Logistic regression (for binary outcomes) and linear regression (for continuous outcomes) were used to estimate the association between each of the 30 PGS and each of the 5 outcomes in independent regression models. For all phenotypes except onset age (for which we excluded age as a predictor), we included age, sex, diagnosis, and the first ten genetic principal components as covariates. Additionally, for admission length and total admissions, we also included year of study entry as an additional covariate.

For each PGS-outcome association tested, we considered two thresholds for statistical significance. Nominal statistical significance was considered at a p-value threshold of 0.05. For a more stringent significance threshold, we applied the Benjamini-Hochberg method within each outcome to calculate false discovery rate (FDR) corrected p-values, considering values below 0.05 to indicate strong evidence of association.

#### Calculation of Variance Explained

We calculated the variance explained in the outcome (*R*^*2*^) by each polygenic predictor as the difference in *R*^*2*^ between the full regression model, which included the polygenic predictor along with other covariates, and a null regression model, which included only covariates. For binary traits, we transformed *R*^*2*^ estimates onto the liability scale using a method which accounts for potential case-control imbalance relative to the population prevalence. For psychosis, we assumed a population prevalence of 0.01, while for self-harm/suicide and aggressive acts, we used the sample prevalence (0.2 for both outcomes) as an approximation of the population prevalence. Confidence intervals for *R*^*2*^ were calculated using the *psychometric* package v2.3 in R (Fletcher, 2022).

## Results

After genomic quality control and imputation, a total of 740 unrelated individuals of Southern Chinese ancestry were available for further analysis (678 cases and 62 controls). Table 1 provides a summary of the five clinical phenotypes measured in patients.

**Table 1.** Summary Statistics of Clinical Phenotypes Measured in Psychosis Patients.

| *Clinical Phenotype* | *N* | *mean* | *sd* | *median* | *min* | *Max* |
| --- | --- | --- | --- | --- | --- | --- |
| Onset Age | 635 | 26.20 | 9.08 | 24 | 12 | 58 |
| Self-Harm/Suicide (binary) | 637 | 0.20 | 0.40 | - | - | - |
| Aggressive Acts (binary) | 637 | 0.21 | 0.41 | - | - | - |
| Length of stay in all past psychiatric hospitalization(s) (days) | 655 | 63.43 | 111.51 | 29 | 0 | 1195 |
| Total number of past psychiatric hospitalization(s) | 678 | 1.13 | 1.27 | 1 | 0 | 8 |

*Note. This table summarizes the final analytic sample characteristics for each of five clinical phenotypes of interest among psychosis cases who passed genotyping quality control filters.*

### Psychosis Risk Prediction

#### Primary Aim

A total of 30 PGS were tested for association with case-control status. The variance explained in case-control status by each PGS is shown in Figure 1. Significant associations at nominal and FDR-corrected thresholds are summarized in Table 2, while complete association results are provided in Supplementary Table 3. The genome-wide PGS for SCZ showed strong evidence for association with case-control status (logOR=0.57, s.e.=0.14, p=8.9e-5, FDR=2.6e-3), explaining 3.7% of phenotypic variance on the liability scale. Four pPGS were also strongly associated with case control status, namely SCZ-weighted pPGS for nervous system development (logOR=0.50, s.e.=0.14, p=2.5e-4, FDR=3.0e-3), regulation of neuron differentiation (logOR=0.50, s.e.=0.14, p=3.0e-4, FDR=3.0e-3), and synapse (logOR=0.40, s.e.=0.13, p=2.8e-3, FDR=2.1e-2), as well as BIP-weighted regulation of neuron differentiation (logOR=0.39, s.e.=0.14, p=5.9e-3, FDR=3.5e-2). These four pPGS each explained between 1.8% and 3.3% of variance in psychosis risk on the liability scale.

**Figure 1.**
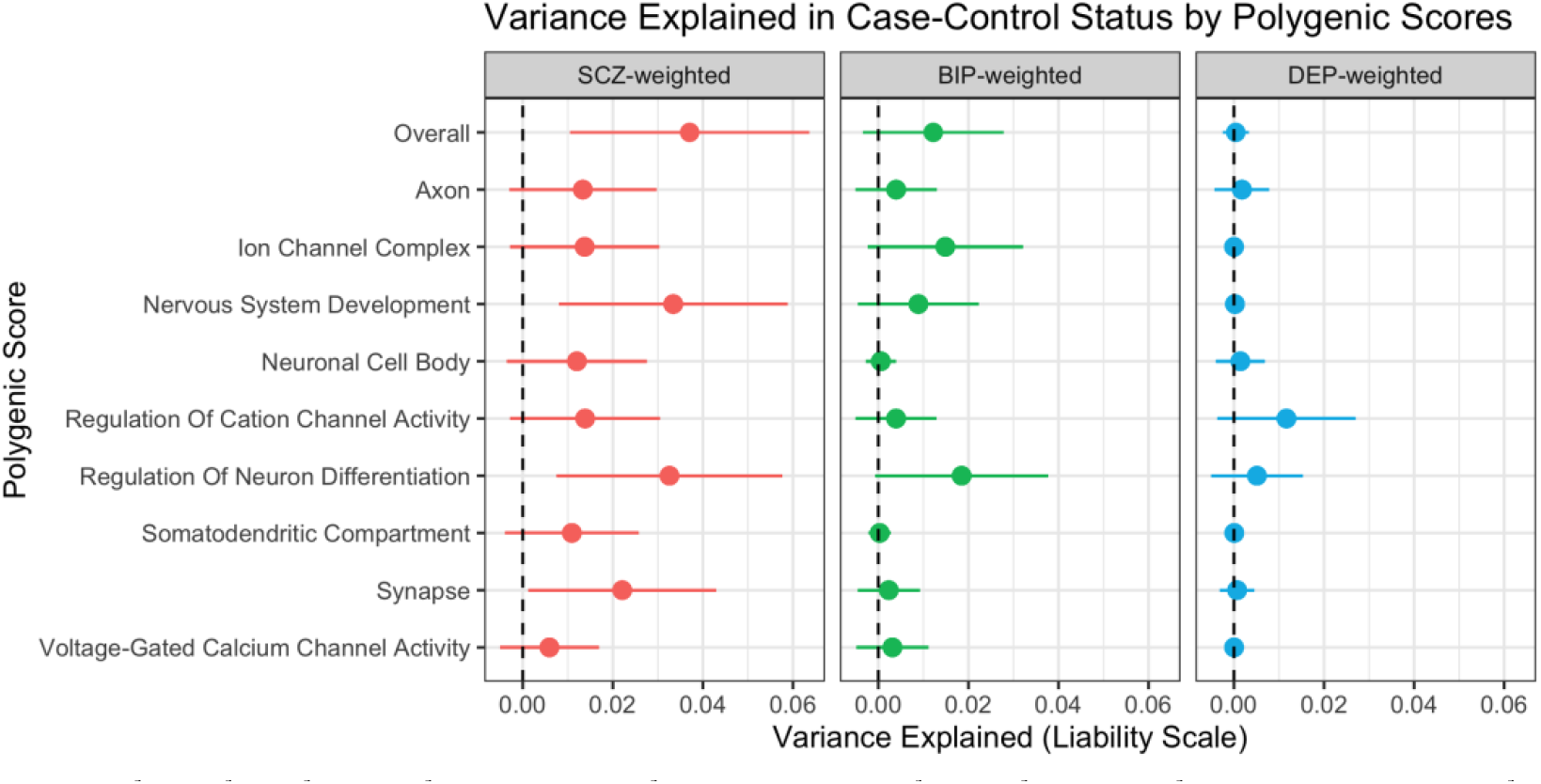
Variance Explained in Case-Control Status by Polygenic Scores. Note. This plot shows the estimated variance explained in psychosis case-control status by overall genome-wide polygenic scores (PGS) and each of nine pathway-specific PGS using weights derived from schizophrenia (SCZ), bipolar disorder (BIP) or depression (DEP) summary statistics. Error bars represent 95% confidence intervals for the variance explained.

**Table 2.** FDR-Corrected and Nominally Significant PGS Associations.

| Outcome | GWAS | N<sub>SNP</sub> | Pathway | R<sup>2</sup> | R<sup>2</sup><sub>liab</sub> | Effect | SE | P | FDR |
| --- | --- | --- | --- | --- | --- | --- | --- | --- | --- |
| Case-Control | SCZ | 659255 | Overall | 0.02 | 0.04 | 0.57 | 0.14 | 8.92e-05 | **2.68e-03** |
|  | SCZ | 11208 | Nervous System Development | 0.02 | 0.03 | 0.50 | 0.14 | 2.49e-04 | **2.96e-03** |
|  | SCZ | 951 | Regulation Of Neuron Differentiation | 0.02 | 0.03 | 0.50 | 0.14 | 2.96e-04 | **2.96e-03** |
|  | SCZ | 8067 | Synapse | 0.01 | 0.02 | 0.40 | 0.13 | 2.78e-03 | **2.08e-02** |
|  | SCZ | 2098 | Ion Channel Complex | 0.01 | 0.01 | 0.34 | 0.14 | 1.66e-02 | 6.35e-02 |
|  | SCZ | 1038 | Regulation Of Cation Channel Activity | 0.01 | 0.01 | 0.32 | 0.14 | 1.82e-02 | 6.35e-02 |
|  | SCZ | 3816 | Axon | 0.01 | 0.01 | 0.32 | 0.14 | 1.90e-02 | 6.35e-02 |
|  | SCZ | 2859 | Neuronal Cell Body | 0.01 | 0.01 | 0.30 | 0.13 | 2.46e-02 | 6.58e-02 |
|  | SCZ | 4806 | Somatodendritic Compartment | 0.01 | 0.01 | 0.29 | 0.13 | 3.40e-02 | 7.85e-02 |
|  | BIP | 1070 | Regulation Of Neuron Differentiation | 0.01 | 0.02 | 0.39 | 0.14 | 5.89e-03 | **3.53e-02** |
|  | BIP | 2463 | Ion Channel Complex | 0.01 | 0.01 | 0.33 | 0.13 | 1.40e-02 | 6.35e-02 |
|  | BIP | 658078 | Overall | 0.01 | 0.01 | 0.30 | 0.13 | 2.37e-02 | 6.58e-02 |
|  | DEP | 1222 | Regulation Of Cation Channel Activity | 0.01 | 0.01 | -0.30 | 0.14 | 2.63e-02 | 6.58e-02 |
| *Number of Admissions* | BIP | 2463 | Ion Channel Complex | 0.01 | -- | 0.16 | 0.05 | 1.48e-03 | **4.43e-02** |
|  | BIP | 620 | Voltage-Gated Calcium Channel Activity | 0.01 | -- | 0.12 | 0.05 | 1.71e-02 | 2.51e-01 |
|  | BIP | 658078 | Overall | 0.01 | -- | 0.11 | 0.05 | 3.27e-02 | 2.51e-01 |
|  | BIP | 1190 | Regulation Of Cation Channel Activity | 0.01 | -- | 0.10 | 0.05 | 4.15e-02 | 2.51e-01 |
|  | BIP | 5505 | Somatodendritic Compartment | 0.01 | -- | 0.10 | 0.05 | 4.76e-02 | 2.51e-01 |
| *Onset Age* | SCZ | 951 | Regulation Of Neuron Differentiation | 0.01 | -- | -0.73 | 0.34 | 3.27e-02 | 3.68e-01 |
|  | BIP | 2463 | Ion Channel Complex | 0.01 | NA | -0.67 | 0.34 | 4.91e-02 | 3.68e-01 |
|  | DEP | 1222 | Regulation Of Cation Channel Activity | 0.01 | NA | -0.82 | 0.34 | 1.69e-02 | 3.68e-01 |
|  | DEP | 644 | Voltage-Gated Calcium Channel Activity | 0.01 | NA | -0.68 | 0.34 | 4.75e-02 | 3.68e-01 |
| *Self-Harm/Suicide* | SCZ | 659255 | Overall | 0.01 | 0.01 | -0.27 | 0.11 | 1.14e-02 | 3.42e-01 |
|  | DEP | 644 | Voltage-Gated Calcium Channel Activity | 0.01 | 0.01 | -0.23 | 0.11 | 3.26e-02 | 4.90e-01 |
| *Total Admission Length* | BIP | 2463 | Ion Channel Complex | 0.01 | -- | 0.17 | 0.08 | 3.45e-02 | 5.33e-01 |
|  | BIP | 1190 | Regulation Of Cation Channel Activity | 0.01 | -- | 0.16 | 0.08 | 4.23e-02 | 5.33e-01 |

*Note. This table shows the associations between polygenic scores (PGS) and case-control status and clinical outcomes among patients. The GWAS column indicates the summary statistics used to weight the pPGS. Pathway indicates which set of genes were included in the PGS calculation, where “Overall” indicates the genome-wide PGS. $N_{\text{SNP}}$ indicates the number of single nucleotide polymorphisms contributing to the PGS. $R^2$ indicates the variance explained on the observed scale, while $R^2_{\text{liab}}$ is the variance explained on the liability scale. FDR indicates the false-discovery rate-corrected p-value, applying the Benjamini-Hochberg method independently within each outcome, correcting for the 30 tests of association with PGS. $\text{FDR} < 0.05$ are indicated in bold. Effect estimates represent log odds-ratio from logistic regression (for binary outcomes) and beta coefficients from linear regression (for continuous outcomes). All PGS were standardized prior to modeling.*

A post-hoc joint analysis including these four pPGS simultaneously as predictors of case-control status indicated that they may still represent overlapping signals, since only the SCZ-weighted nervous system development and regulation of neuron differentiation pPGS remained significant at p<0.05 in the joint model. Modeled together, these two pPGS explained 5.2% of variance in psychosis liability, compared to 3.7% accounted for by the genome-wide SCZ PGS. Interestingly, both pPGS remained significant at p < 0.05 even when jointly modeled together with the overall genome-wide SCZ PGS. The addition of the genome-wide SCZ PGS accounted for an increase of 1.6% variance explained in psychosis liability on top of the variance explained by the two pPGS alone. Figure 2 shows the proportion of cases across PGS quantiles for each of on these three PGS.

**Figure 2.**
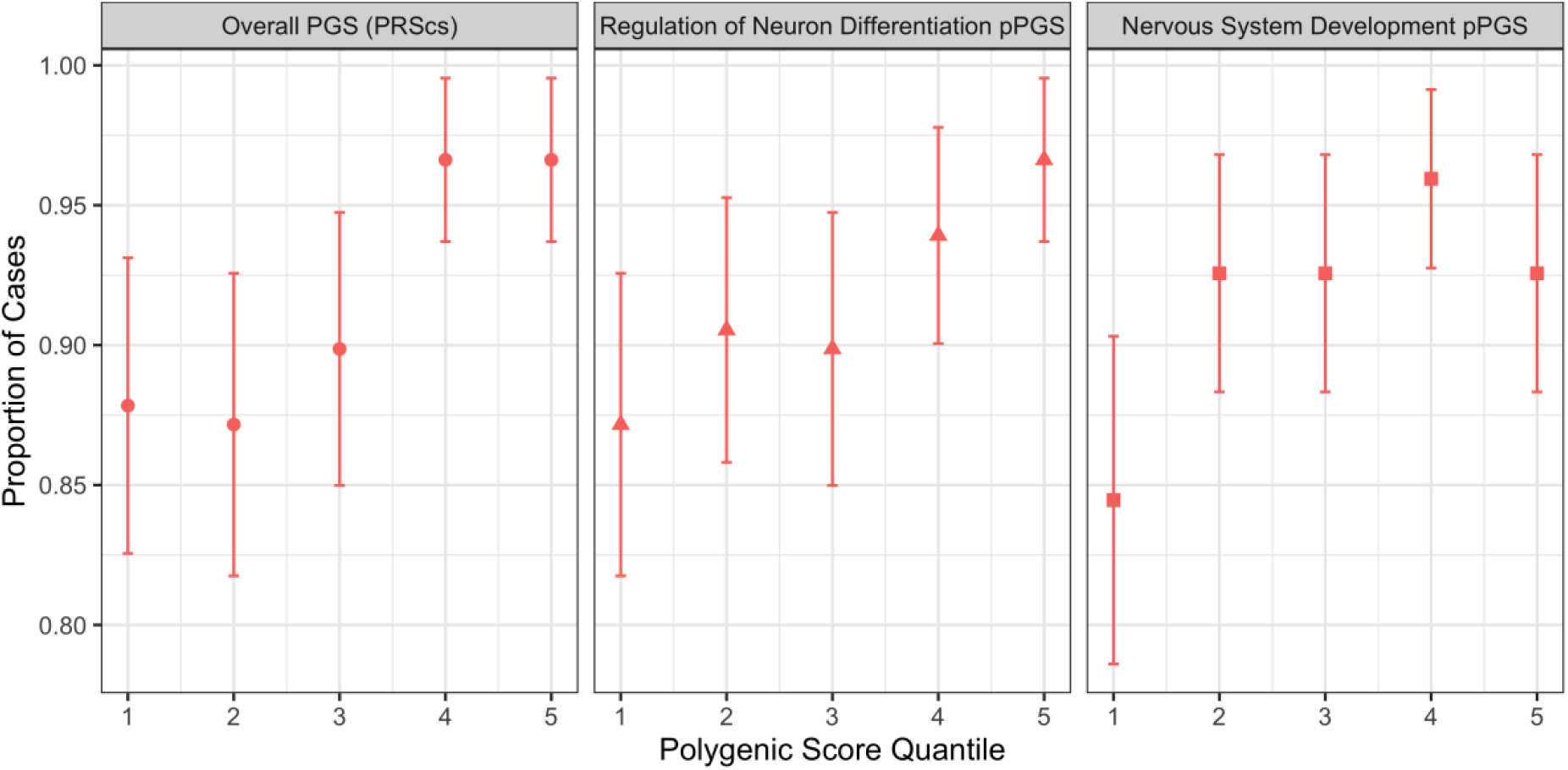
Proportion of Cases Across SCZ PGS and pPGS Quintiles. Note. This figure shows the proportion of psychosis cases across increasing schizophrenia-weighted polygenic score (PGS) quantiles for an overall genome-wide PGS, and two pathway-specific PGS (pPGS) defined by genes involved in regulation of neuron differentiation and nervous system development.

Eight additional PGS showed nominally significant associations with case-control status, each independently explaining approximately 1% of variance in psychosis liability. These included 5 SCZ-weighted pPGS, the overall BIP PGS, a BIP-weighted pPGS, and a DEP-weighted pPGS.

### Clinical Outcome Prediction

#### Secondary Aim

Each of the 30 PGS were also tested for association with each of five clinical outcomes. Statistically significant associations are reported in Table 2, while all results are presented in Supplementary Table 3. Only one clinical phenotype showed strong evidence for significant association with a PGS after FDR correction. Namely, the BIP-weighted ion channel complex pPGS explained 1.4% of the variance in total number of inpatient admissions. The association between this pPGS and total admissions is shown alongside the overall genome-wide BIP PGS in Figure 3. Post-hoc joint modeling of the overall BIP PGS together with the BIP ion channel complex pPGS suggested that a large proportion of overall genetic risk may be accounted for by the ion channel complex pathway, given that the overall BIP PGS was not significant when modeled jointly and only accounted for 0.3% of additional variance on top of that explained by the ion channel complex pPGS.

**Figure 3.**
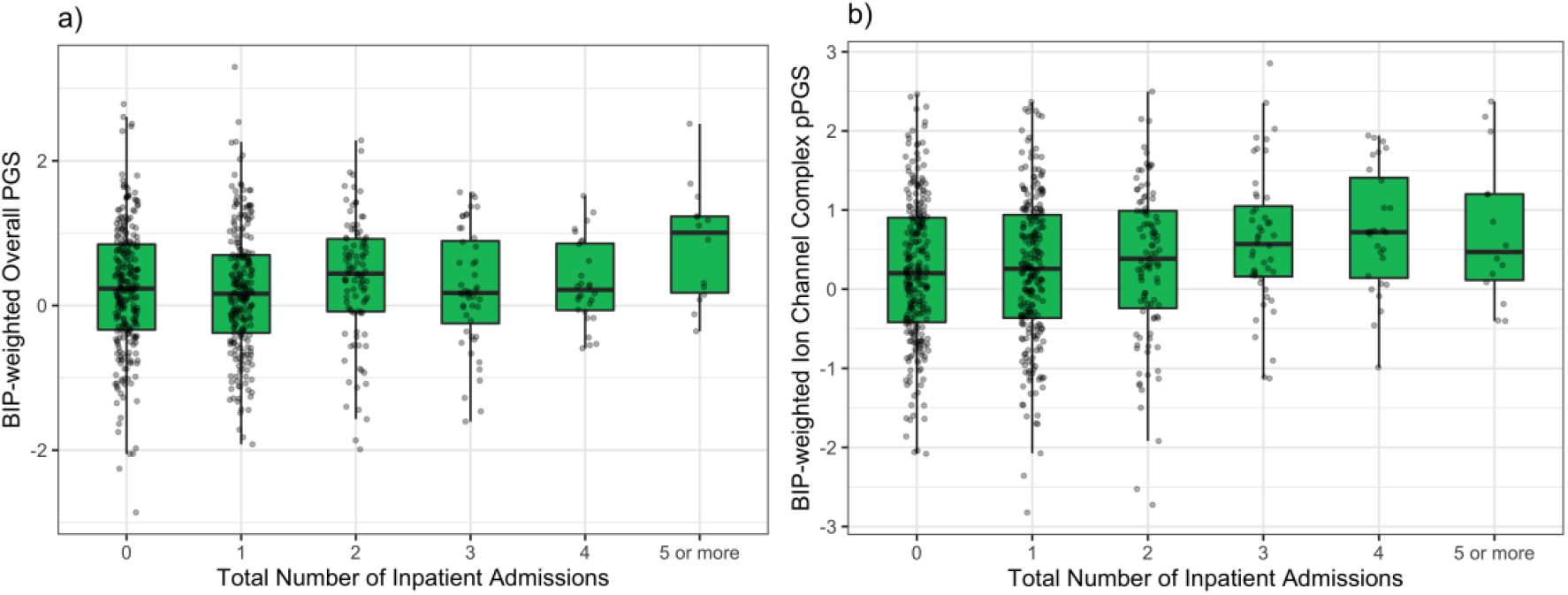
Association of Bipolar-Weighted PGS and Total Number of Inpatient Admissions. Note. this plot shows the association between total number of inpatient admissions and overall genome-wide polygenic scores (PGS) and Ion Channel Complex pathway PGS (pPGS), both using weights derived from bipolar disorder (BIP) summary statistics.

As shown in Table 2, 12 nominally significant associations were identified between SCZ-BIP- and DEP-weighted PGS and the clinical outcomes. These included associations with onset age, self-harm/suicide, admission length, and total admissions. However, these findings require replication in datasets with greater power.

## Discussion

In a sample of psychosis patients and controls, this study examined associations between polygenic scores, risk for psychosis, and a number of important clinical outcomes, utilizing both traditional genome-wide PGS and pathway-specific pPGS. Strong evidence supported an association between higher genome-wide SCZ PGS and increased risk of psychosis, with significant contributions also accounted for by both nervous system development and regulation of neuron differentiation pPGS. The BIP-weighted ion channel complex pPGS was strongly associated with increased number of total inpatient psychiatric admissions. This pathway appears to account for a major portion of overall BIP PGS signal.

The variance in psychosis liability explained by the genome-wide SCZ PGS in this study (3.7%) is similar to estimates from other East Asian samples (Ikeda et al., 2019; Lam et al., 2019), but is significantly lower than the performance of SCZ PGS in European samples, which has been estimated to be around 7% (Trubetskoy et al., 2022). However, joint modeling of the SCZ-weighted overall PGS, nervous system development pPGS, and regulation of neuron differentiation pPGS together accounted for a total of 6.9% of variance in psychosis liability in the current sample, approaching the predictive power of PGS in European ancestry samples.

One potential explanation is that modeling individual variation in specific schizophrenia-related pathways through pPGS may capture additional signal that is lost within the large proportion of random noise contained in the overall genome-wide PGS. Thus, incorporating pathway information into polygenic risk models, essentially allowing for secondary weighting of biological pathways, has the potential to increase predictive power, even across ancestral populations.

Although PGS and pPGS have largely been used in previous studies to predict liability to disease, they can also be used to predict important clinical outcomes among patients. This study found evidence for an association between BIP PGS and number of inpatient admissions among psychosis patients, which was largely driven by the ion channel complex pPGS. Although BIP diagnosis was nominally associated with a higher number of inpatient admissions, the pPGS association remained significant while controlling for diagnosis as a jointly modeled covariate. Nominally significant BIP pPGS associations with total admissions were also observed for the voltage-gated calcium channel activity and regulation of cation channel activity pathways. Prior genetic studies have linked ion channel pathway genes to risk for schizophrenia and bipolar disorder (Harrison et al., 2018; Trubetskoy et al., 2022), but no prior study to our knowledge has reported a link between ion channel pathway variants and increased risk for inpatient admissions. These results suggest that BIP-associated risk localized to the ion channel complex pathway may be especially important in predicting psychosis severity, specifically increased risk of inpatient admissions, but requires replication and follow-up studies to understand possible mechanisms. For example, future studies may examine additional factors associated with this pPGS through a phenome-wide association study to provide clues about potential mediating factors, while brain imaging studies may examine whether this pPGS associates with specific differences in brain morphology.

Despite encouraging results supporting the utility of pPGS in improving cross-ancestry prediction of complex traits, our study still suffers a few limitations. Most notably, our sample size is relatively small, requiring replication in larger samples. Secondly, the method employed for calculating pPGS employs a relatively simple scoring algorithm, which is currently necessary to ensure complete coverage of pathway-specific markers. Future work should focus on adapting more sophisticated and powerful algorithms, such as PRS-cs (Ge et al., 2019). Nonetheless, one of the major strengths of this study is its sample, which includes psychosis cases with deep phenotyping and non-psychiatric controls from an under-studied population in genetic research.

This study demonstrates the utility of pPGS for improving prediction of liability to psychosis in a Chinese sample, approaching the variance explained by genome-wide PGS in European samples. Furthermore, pPGS provide greater insight into mechanisms underlying genetic risk for disease and clinical outcomes, demonstrated by the association of multiple SCZ pPGS with case-control status, and of BIP ion channel complex pPGS and a greater number of inpatient psychiatric admissions.

## Data Availability

As stipulated in the ethical approval for this study, the data are confidential and thus are not publicly available.

