## Supplementary Tables 1 & 2 for "Pathway-Specific Polygenic Scores Improve Cross-Ancestry Prediction of Psychosis and Clinical Outcomes" (from another version of this preprint: version 1, 2023-09-02)

**Supplementary Table 1: Diagnostic composition of our sample**

| Subjects | Diagnosis | N |
| --- | --- | --- |
| Clinical participants | Schizophrenia | 483 |
|  | Bipolar Disorder | 56 |
|  | Acute Transient Psychotic Disorder | 28 |
|  | Schizoaffective Disorder | 26 |
|  | Delusional Disorder | 20 |
|  | Psychosis not otherwise specified | 65 |
| Controls | - | 62 |

**Supplementary Table 2: Operational definitions of the six phenotypic variables**

| Variable | Nature | Operational definitions |
| --- | --- | --- |
| Case-control status | Binary | Best-possible end-point estimate. Schizophrenia-spectrum disorders included the diagnoses of schizophrenia, bipolar disorder, delusional disorder, schizoaffective disorder, acute and transient psychotic disorder, and psychosis not otherwise specified. |
| Onset age | Continuous | The onset age of first frank psychotic symptoms documented at computerized medical record. |
| History of self-harm/suicide behaviors | Binary | We defined subjects as having history of self-harm/suicide behaviors if the medical records documented the followings: suicide attempt of any kind, suicide gesture, deliberate self-harm, drug overdose. We did not include “suicidal ideation” (without acting out). |
| History of aggressive acts | Binary | We defined subjects as having “history of aggressive acts” if the medical records documented the followings: aggressive acts, violent acts, assaults, attacking others. The aggressive acts could target at people or objects. The aggressive acts did not necessarily lead to psychiatric hospitalizations or Police custody or criminal records. |
| Length of stay in all past psychiatric hospitalization(s) (days) | Continuous | We identified the admission and discharge date of all past psychiatric hospitalization(s), and calculated the total number of calendar days of inpatient stay for each participants. The computerized medical record of the Hospital Authority Hong Kong has completely recorded all inpatient treatment records in our sample. |
| Total number of past psychiatric hospitalization(s) | Continuous | We counted the number of past psychiatric hospitalization(s). The computerized medical record of the Hospital Authority Hong Kong has completely recorded all inpatient treatment records in our sample. |
